# Predicting long-term prognosis after percutaneous coronary intervention in patients with acute coronary syndromes: a prospective nested case-control analysis for county-level health services

**DOI:** 10.1101/2023.07.05.23292269

**Authors:** Yue Lu, Yaqian Wang, Bo Zhou

**Affiliations:** Department of Clinical Epidemiology and Evidence-Based Medicine, The First Hospital of China Medical University, Shenyang 110000, Liaoning, China

**Author notes:** Correspondence: Bo Zhou. contributed equally to this work and share first authorship (Yue Lu, Yaqian Wang).

**Keywords:** ACS, PCI, MACEs, predictive nomogram

## Abstract

**Objective:** We aimed to establish and authenticate a clinical prognostic nomogram for predicting long-term Major Adverse Cardiovascular Events (MACEs) among high-risk patients who have undergone Percutaneous Coronary Intervention (PCI) in county-level health service.

**Methods:** This prospective study included Acute Coronary Syndrome (ACS) patients treated with PCI at six county-level hospitals between September 2018 and August 2019, selected from both the original and external validation cohorts. Least Absolute Shrinkage and Selection Operator (LASSO) regression techniques and logistic regression were used to assess potential risk factors and construct a risk predictive nomogram. Additionally, the potential non-linear relationships between continuous variables were tested using Restricted Cubic Splines (RCS). The performance of the nomogram was evaluated based on the Receiver Operating Characteristic (ROC) curve analysis, Calibration Curve, Decision Curve Analysis (DCA), and Clinical Impact Curve (CIC).

**Results:** The original and external validation cohorts comprised 520 and 1061 patients, respectively. The final nomogram was developed using nine clinical variables: Age, Killip functional classification III-IV, Hypertension, Hyperhomocysteinemia, Heart failure, Number of stents, Multivessel disease, Low-density Lipoprotein Cholesterol, and Left Ventricular Ejection Fraction. The AUC of the nomogram was 0.79 and 0.75 in the original and validation cohorts, respectively. The DCA and CIC validated the clinical value of the constructed prognostic nomogram.

**Conclusion:** Herein, we developed and validated a prognostic nomogram for predicting the probability of 3-year MACEs in ACS patients who underwent PCI at county-level hospitals. The nomogram could provide a precise risk assessment for secondary prevention in ACS patients receiving PCI.

## Background

Coronary Heart Disease (CHD) is a major contributor to global health and socioeconomic burdens, especially with the rising aging of the world population. As an alarming emergence, CHD is a global public health issue posing a severe threat to human health [1]. Despite an increase in the overall CHD burden, age-adjusted CHD mortality is decreasing in developed countries[2, 3]. Low- and Middle-Income Countries (LMIC) face a substantial cardiovascular disease burden as urbanization and societal transformation escalate. According to the Report on Cardiovascular Health and Diseases in China data from 2019, rural and urban areas experienced a significant burden of mortality due to cardiovascular disease (CVD), with CVD accounting for 46.74% and 44.26% of all deaths in these respective regions[4]. Furthermore, CHD severity appears to follow a reverse socioeconomic gradient in developing nations[5]. A prospective analytical study conducted in India discovered notable socioeconomic disparities in access to primary and secondary prevention for Acute Coronary Syndrome (ACS) management[6].

As a life-threatening symptom of coronary heart disease, ACS includes ST-segment Elevation Myocardial Infarction (STEMI), Non-ST-segment Elevation Myocardial Infarction (NSTEMI), and Unstable Angina (UA)[7, 8]. A combination of oral medications and surgical procedures are commonly used to manage patients with ACS [9, 10]. Percutaneous Coronary Intervention (PCI), which involves balloon dilatation and stenting, is the preferred method of reperfusion therapy for ACS patients[10–12]. Despite undergoing PCI, a subset of ACS patients continue to experience Major Adverse Cardiovascular Events (MACEs), including non-fatal myocardial infarction, non-fatal ischemic stroke, death, and bleeding events[13].

Although the prediction models for clinical assessment of ACS patients in primary and secondary care post-PCI exhibit high accuracy, their clinical applicability is limited due to their simplicity and inconsistency[14–16]. Furthermore, the Human Development Index (HDI) is strongly associated with CHD prevalence in developing countries, and the current allocation of healthcare resources to county-level hospitals in LMIC is relatively low. Therefore, to ensure the continued enhancement of quality healthcare services, there is an urgent need to further develop patient prognostic assessment methods [17, 18].

Consequently, our study aimed to validate a prognostic model capable of predicting the likelihood of experiencing MACEs in ACS patients who underwent PCI at a county-level hospitals. We employed a multicenter external validation strategy to serve as a reference for the swift screening of high-risk individuals and early clinical intervention. Moreover, our study aimed to foster the advancement of quality healthcare in regions with limited healthcare resources.

## Methods

### Study Population and Design

From September 2018 to August 2019, a prospective nested case-control study was undertaken on a primary cohort of ACS patients enrolled for PCI at six county-level hospitals within Liaoning Province, China. Across all study centers, 1795 individuals were diagnosed with ACS and treated with PCI based on their clinical conditions. A total of 1741 patients were followed up from recruitment to August 2022, and 3.0% of cases (n = 54) were removed due to incomplete clinical information and exclusion criteria. A total of 1581 patients were finally analyzed after three years of follow-up. The study enrolment and results are shown in Figure 1. The original cohort used to build the prognostic assessment model consisted of three centers (n=520) matched considering their socioeconomic status and environmental conditions, and three other cohorts (n=1061) were used to validate the model prospectively. The case group consisted of 143 ACS patients who experienced a major adverse cardiovascular event during the follow-up period from September 2018 to August 2019. On the other hand, the control group included patients who did not experience an adverse cardiovascular event until the end of the follow-up period.

**Figure 1.**
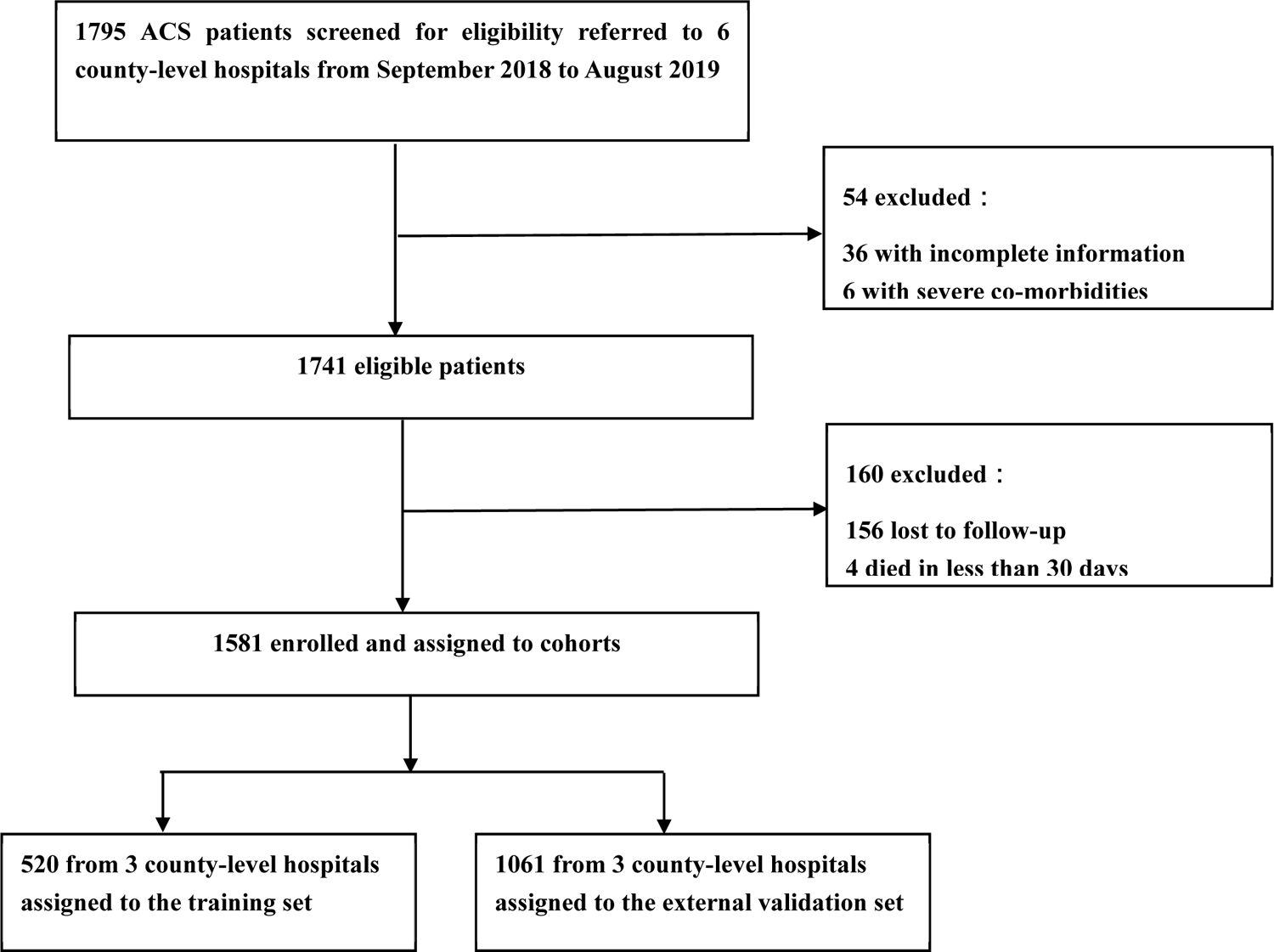
Study flow diagram.

The following were the inclusion criteria: (1) Patients with ACS who met the Chinese Medical Association’s guidelines for diagnosis and treatment of acute coronary syndromes (Supplementary methods contain definitions for each subset of ACS); (2) Patients who underwent complete revascularization post-PCI. The exclusion criteria were as follows: (1) Patients with hematologic disease, multiple organ failure, or a cancer diagnosis; (2) Patients with significant comorbidity, trauma, or surgery; (3) Patients with insufficient clinical or laboratory data; (4) Patients who died within 30 days.

The study was approved by the institutional review board of the First Hospital of China Medical University (approval number: [2019]189). Written informed consent was obtained from all surviving participants or the next of kin who provided information about the deceased participants.

The study adheres to the Transparent Reporting of a Multivariable Prediction Model for Individual Prognosis or Diagnosis (TRIPOD) reporting guidelines[19].

### Clinical Endpoint and Definitions

The primary endpoint of this study was MACEs, defined as a composite of stroke, heart failure, target lesion revascularization, recurrent myocardial infarction, and all-cause death.

### Data Collection and Follow-up

Baseline characteristics (patient demographic data, medical history, preoperative clinical characteristics, coronary angiography features, laboratory indicators, echocardiography indices, and medication use during hospitalization) of the external and original cohorts are shown in STable 1.

Through telephone (to the patients themselves or their relatives), patients were followed up in the first, second, and third years post-PCI, with no further follow-up if a death event was recorded. During follow-up, medication use, daily behavioral habits, and outcome events, including all-cause death, target lesion revascularization, recurrent myocardial infarction, stroke, heart failure, rehospitalization for cardiac reasons, and major bleeding events, were all recorded. The general condition of the patients and the medications taken during the follow-up period are depicted in STable 2.

### Statistical analysis

Normally distributed variables were summarized as means and standard deviations, medians and interquartile ranges represented skewed distributional data, and frequencies or proportions were used to describe categorical variables. The t-test, the rank-sum test, and the chi-square test were applied to compare continuous variables of normal distribution, continuous variables of non-normal distribution, and categorical variables, respectively.

As candidate predictors, 97 clinical features with at least 70% data completeness were evaluated. For the missing values, multiple imputation was performed using random forest. The most useful predictors were filtered using the Least Absolute Shrinkage and Selection Operator (LASSO) regression, which was additionally augmented with 10-fold cross-validation for internal validation. The most predictive covariates were selected by lambda.1se. The predictor factors discovered by the LASSO regression analysis were incorporated using the multivariate logistic regression model. Subsequently, the predictor variables that consistently achieved statistical significance were used to generate the risk score and were represented by the nomogram.

Furthermore, Restricted Cubic Splines (RCS) were used to analyze the association of continuous variables with MACEs incidence among ACS patients post-PCI. The reference values (OR=1) were set at the 10th percentiles, and four knots were placed at the 5th, 35th, 65th, and 95th percentiles of the distribution, respectively. Following that, continuous variables that reported non-linear association were transformed based on RCS and clinical experience to develop an improved predictive model. All statistical analyses were performed using SPSS (version 26.0) and R software (Version R-4.1.3).

## Results

### Characteristics of patients and outcome

After excluding those who were lost to follow-up or had missing data, the study comprised 1581 ACS patients who underwent PCI. The training and validation sets included 520 and 1061 patients, respectively. The baseline and follow-up characteristics of the patients in the training and validation sets are respectively displayed in STable 1 and STable 2. During follow-up, MACEs were detected in 143 (27.5%) but not in 377 training data set instances and in 230 (21.7%) but not in 831 validation set cases.

### Predictor Selection

The LASSO regression included 97 factors evaluated at admission and follow-up (STable 1 and Stable 2 in the Supplement). Twenty-one variables (Age, Killip Functional Classification III-IV (Killip III-IV), Hypertension, dyslipidemia, Hyperhomocysteinemia (HHcy), Heart Failure (HF), Chronotropic Incompetence (CI), Pulmonary Heart Disease (PHD), Number of stents, Multivessel disease, New-Onset Diabetes after PCI (NODAP), Aspartate Aminotransferase (AST), Low-Density Lipoprotein Cholesterol (LDL-C), estimated Glomerular Filtration Rate (eGFR), Total Cholesterol (TC), Blood Urea Nitrogen (BUN), Absolute Neutrophil Count (ANC), Left Ventricular Ejection Fraction (LVEF), Left Atrial Diameter (LAD), Maximum Dilation Pressure (MDP), P_2_Y_12_ Receptor Antagonist (P_2_Y_12_-RA) use after PCI were found to continue to be significant predictors of MACEs (Figure 2).

**Figure 2.**
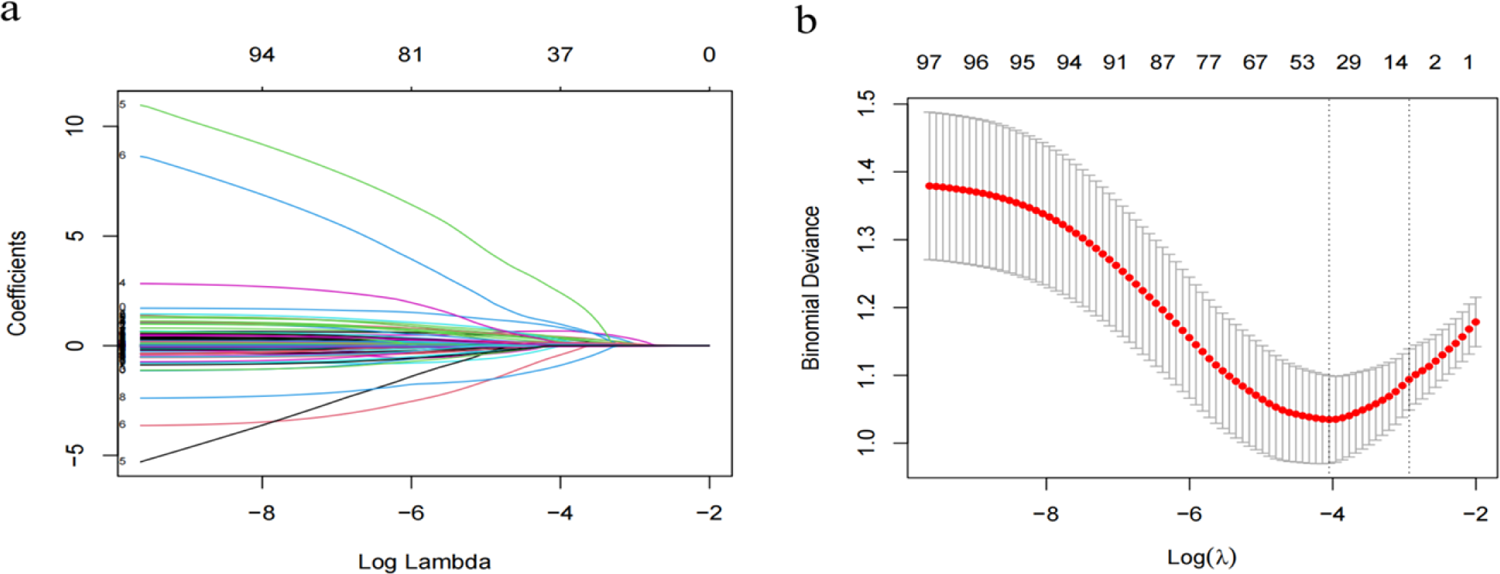
Feature selection using the LASSO binary logistic regression model **Notes**: (**a**) The figure shows the LASSO coefficient curves for 97 variables based on log(lambda). The distribution of the coefficients was generated by the sequence. (**b**) A vertical line was drawn at the value selected using ten-fold cross-validation. The optimal model was acquired when the lambda was 0.03340004, where the optimal lambda resulted in fifteen features with nonzero coefficients.

### Association between continuous variables and predicted outcomes

The correlation between continuous variables selected through LASSO regression and the anticipated outcome was analyzed before developing the prognostic program. We employed restricted cubic splines (RCS) to graphically represent non-linear associations (STable 3). In both the training and external validation sets, the variable LVEF showed a non-linear association with the predicted outcome MACEs (Fig. 3).

**Figure 3.**
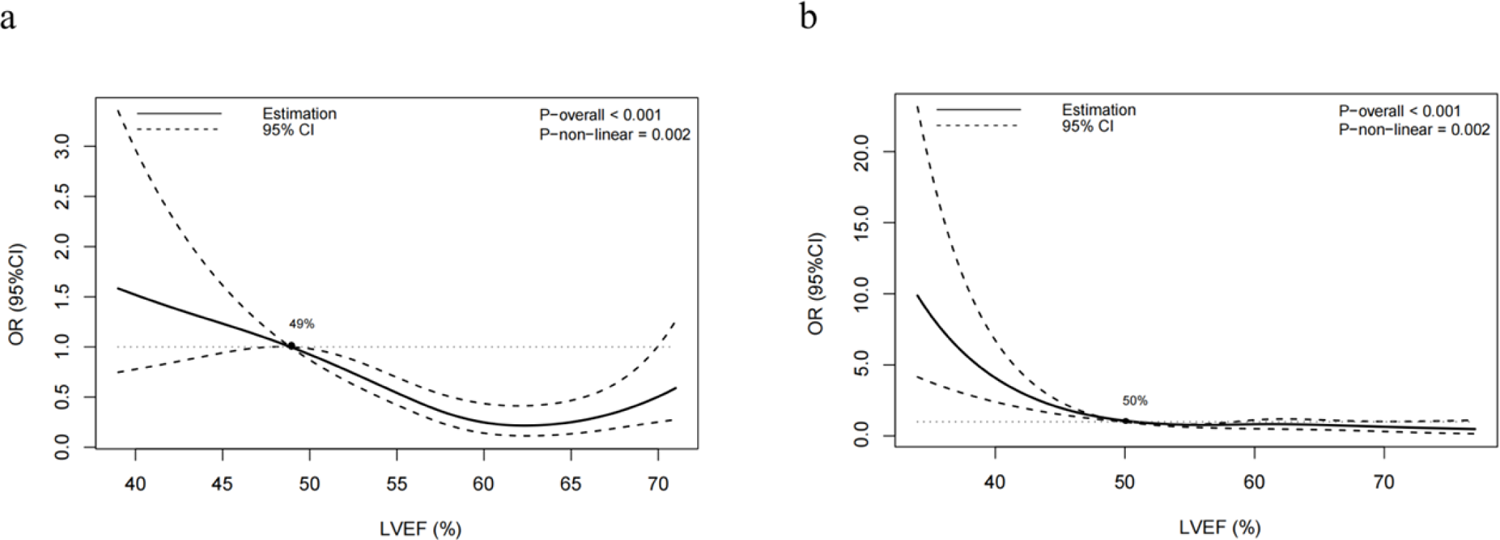
Association of LVEF with risk of MACEs (**a** shows an association of LVEF with risk of MACEs after adjustment for other confounders in the training set; **b** shows an association of LVEF with risk of MACEs after adjustment for other confounders in the external validation set.)

### Development of the multivariate prognostic nomogram

According to the univariate and multivariate logistic regression analysis (P<0.05) (Table 1), 9 out of 21 prospective clinical factors were independently statistically significant predictors of MACEs in the training data set and were incorporated in the prognostic nomogram (Fig. 4). These variables included Age (OR, 1.08; 95%CI, 1.06-1.11; P < 0.001), Killip III-IV (OR, 4.65; 95%CI, 1.43-15.14; P = 0.011), Hypertension (OR, 1.81; 95%CI, 1.11-2.97; P =0.018), HHcy (OR, 4.99; 95%CI, 1.46-17.04; P = 0.010), HF (OR, 1.83; 95%CI, 1.02-3.26; P = 0.042), Number of stents (OR, 1.40; 95%CI, 1.09-1.80; P = 0.008), Multivessel disease (OR, 1.78; 95%CI, 1.13-2.83; P = 0.014), LDLC (OR, 1.54; 95%CI, 1.22-1.93; P < 0.001), and LVEF (OR, 0.26; 95%CI, 0.13-0.49; P < 0.001). A straight line drawn from the point axis upward connected each predictor in a prognostic nomogram to a specific point. The “Total Points” axis was used to display the sum of the scores for each variable. The plotted “Total Points” axis was subsequently connected directly to the probability axis by a vertical line to determine the probability of MACEs.

**Figure 4.**
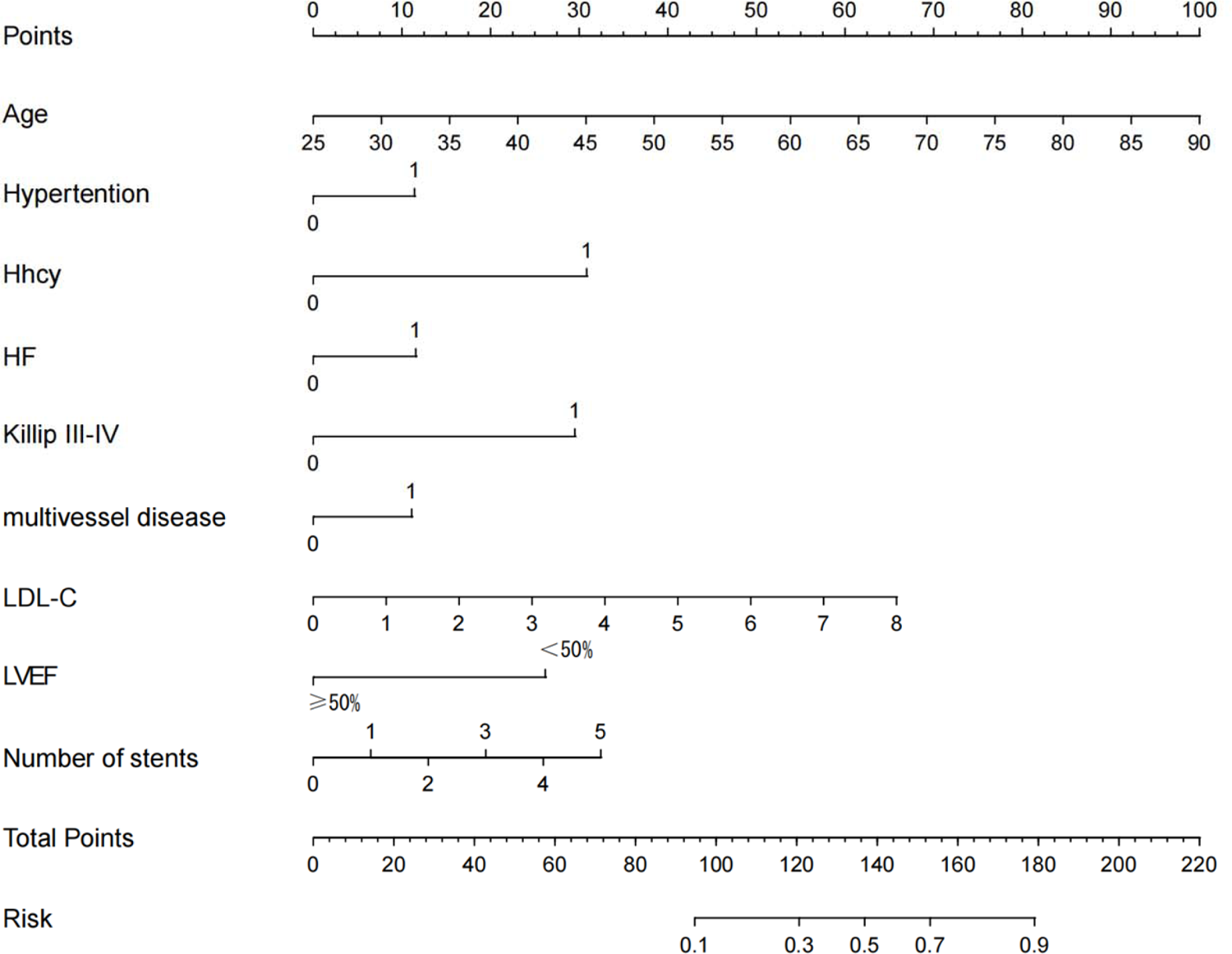
Nomogram used for predicting MACEs after PCI in ACS patients. The final score is calculated as the sum of the individual scores of each of the nine variables included in the nomogram.

**Table 1.**
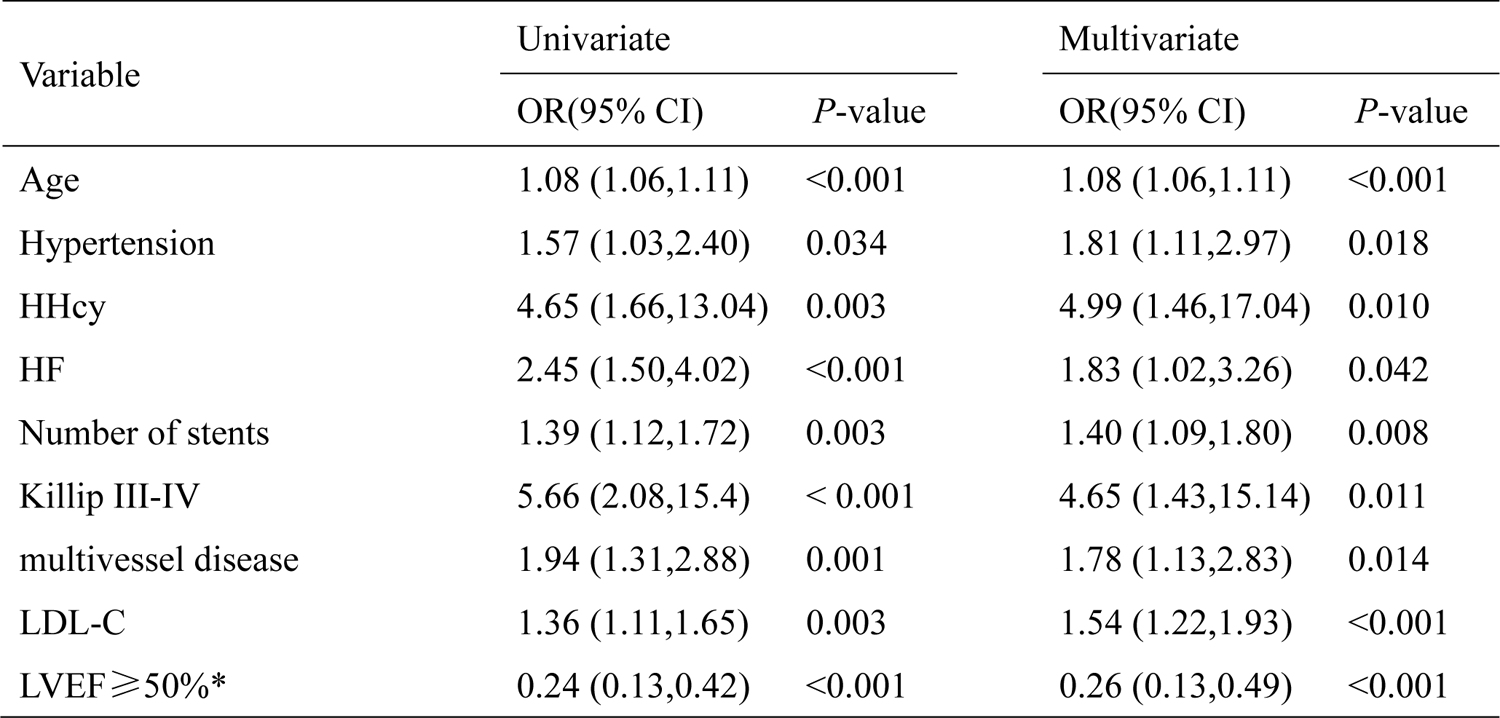
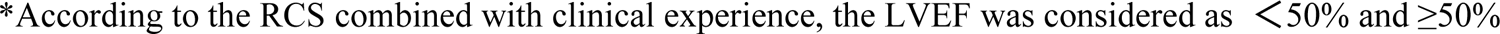
Univariate and multivariable logistics regression analysis of predictive variables in the training set

| Variable | Univariate |  | Multivariate |  |
| --- | --- | --- | --- | --- |
|  | OR(95% CI) | *P*-value | OR(95% CI) | *P*-value |
| Age | 1.08 (1.06,1.11) | <0.001 | 1.08 (1.06,1.11) | <0.001 |
| Hypertension | 1.57 (1.03,2.40) | 0.034 | 1.81 (1.11,2.97) | 0.018 |
| HHcy | 4.65 (1.66,13.04) | 0.003 | 4.99 (1.46,17.04) | 0.010 |
| HF | 2.45 (1.50,4.02) | <0.001 | 1.83 (1.02,3.26) | 0.042 |
| Number of stents | 1.39 (1.12,1.72) | 0.003 | 1.40 (1.09,1.80) | 0.008 |
| Killip III-IV | 5.66 (2.08,15.4) | < 0.001 | 4.65 (1.43,15.14) | 0.011 |
| multivessel disease | 1.94 (1.31,2.88) | 0.001 | 1.78 (1.13,2.83) | 0.014 |
| LDL-C | 1.36 (1.11,1.65) | 0.003 | 1.54 (1.22,1.93) | <0.001 |
| LVEF≥50%* | 0.24 (0.13,0.42) | <0.001 | 0.26 (0.13,0.49) | <0.001 |

\*According to the RCS combined with clinical experience, the LVEF was considered as $<50\%$ and $\geq 50\%$

### Performance of the prognostic nomogram

The Area Under the ROC Curve (AUC) for the model in the training set was 0.79 (95% CI 0.75-0.84), indicating excellent discrimination (Fig 5a). The model’s cutoff value was 0.30, and based on the optimal cutoff points, the sensitivity and specificity were 76.90% and 69.50%, respectively. The calibration plot for the risk of MACEs demonstrated a substantial agreement between nomogram prediction and actual observation (Fig 5b).

**Figure 5.**
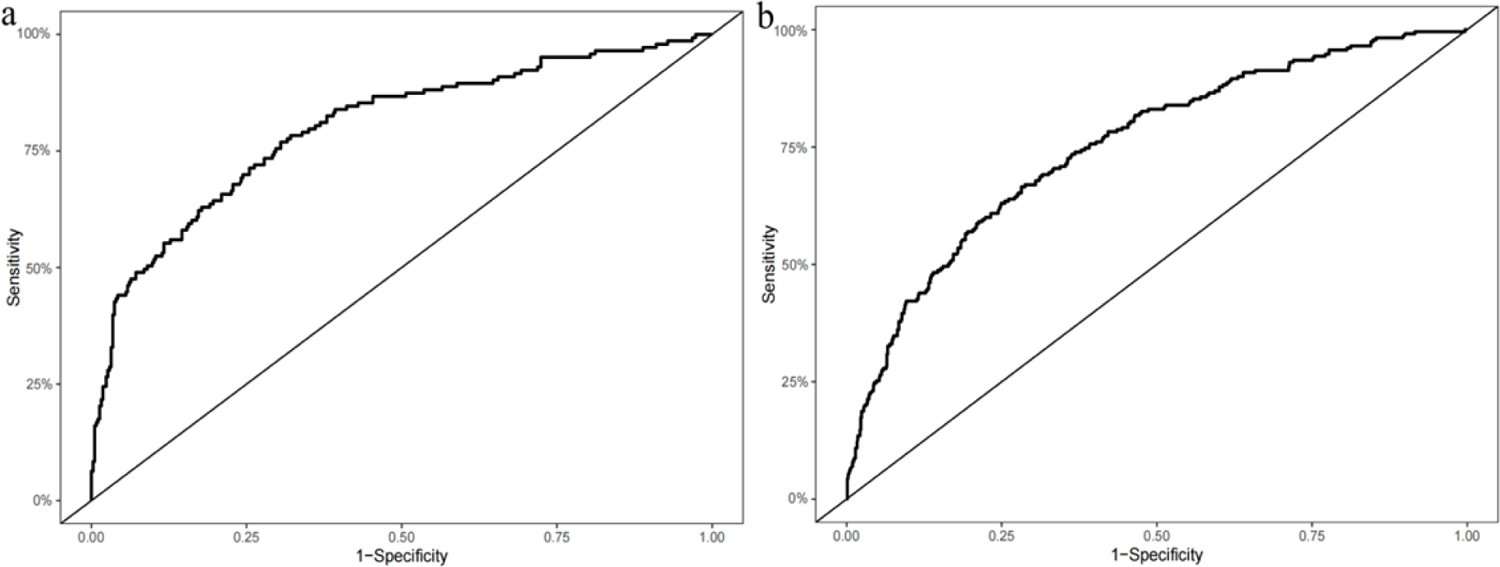
ROC curve of the nomogram for predicting the risk of MACEs after PCI in ACS patients. (a) ROC curve in the training set; (b) ROC curve in the validation set.

### Validation of the prognostic nomogram

In the validation set, the nomogram demonstrated an excellent predictive ability for forecasting the risk of MACEs post-PCI. According to the ROC curve, the AUC value of the model was 0.75 (95% CI: 0.71-0.78) (Fig. 6a), and with these results, the validation set likewise confirmed the nomogram’s favorable calibration (Fig. 6b).

**Figure 6.**
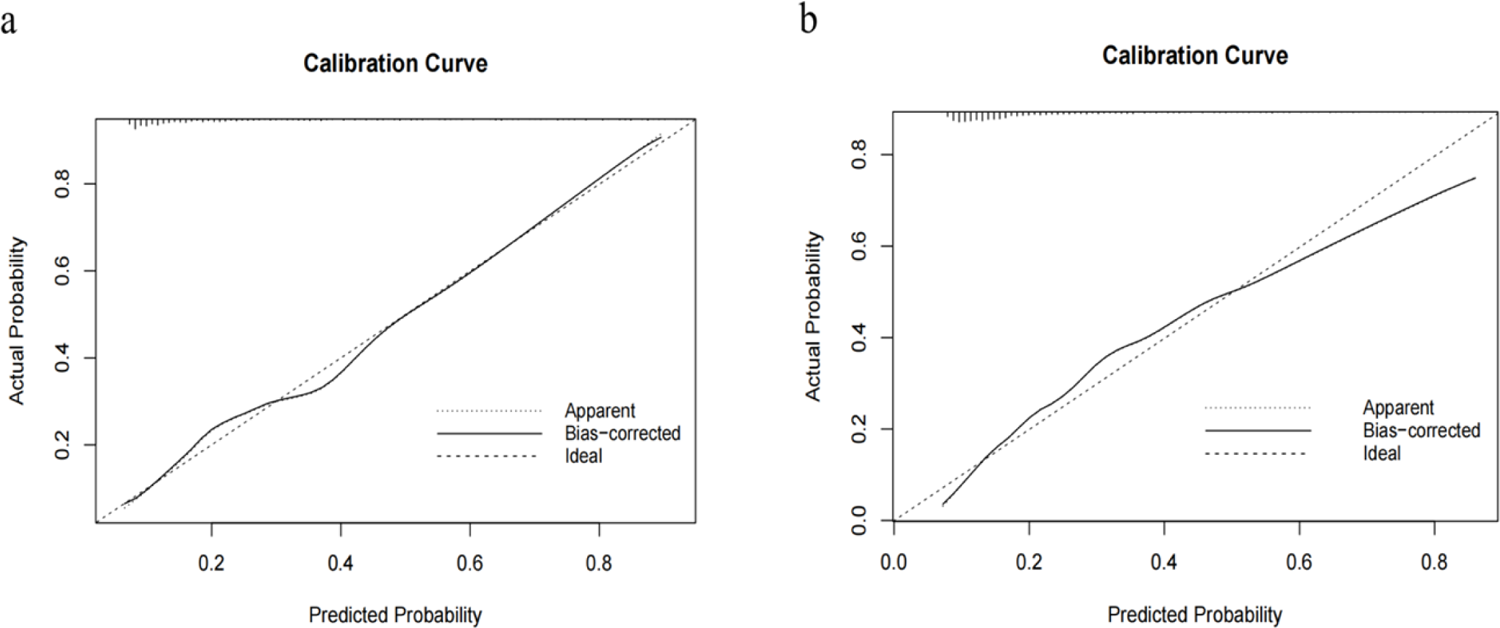
Calibration curve of the nomogram for the training set (a) and the validation set (b). The X-axis represents the overall predicted probability of MACEs after PCI, and the Y-axis represents the actual probability. Model calibration is indicated by the degree of fitting of the curve and the diagonal.

### Clinical Utility

Furthermore, the clinical validity of the nomogram regarding its clinical utility was evaluated using a Decision Curve Analysis (DCA) and Clinical Impact Curve (CIC) (Figures 7 and 8). The DCA demonstrated that the net benefit of using the nomogram outweighed the benefits of the “Treat None” and “Treat All” protocols when the threshold probability of MACEs post-PCI in ACS patients was between 0.00 and 0.8, implying that the nomogram has a promising clinical applicability and applies to both the training and validation sets. The horizontal position of CIC represents the probability threshold, while its vertical coordinate represents the number of people. The blue line represents number of people the model considers to be at high risk of experiencing an adverse event at various probability thresholds. In contrast, the red line shows the proportion of people the model deems to be at high risk at specific probability levels. In conclusion, these findings supported the clinical applicability and accuracy of the nomogram in predicting the probability of MACEs in ACS patients post-PCI.

**Figure 7.**
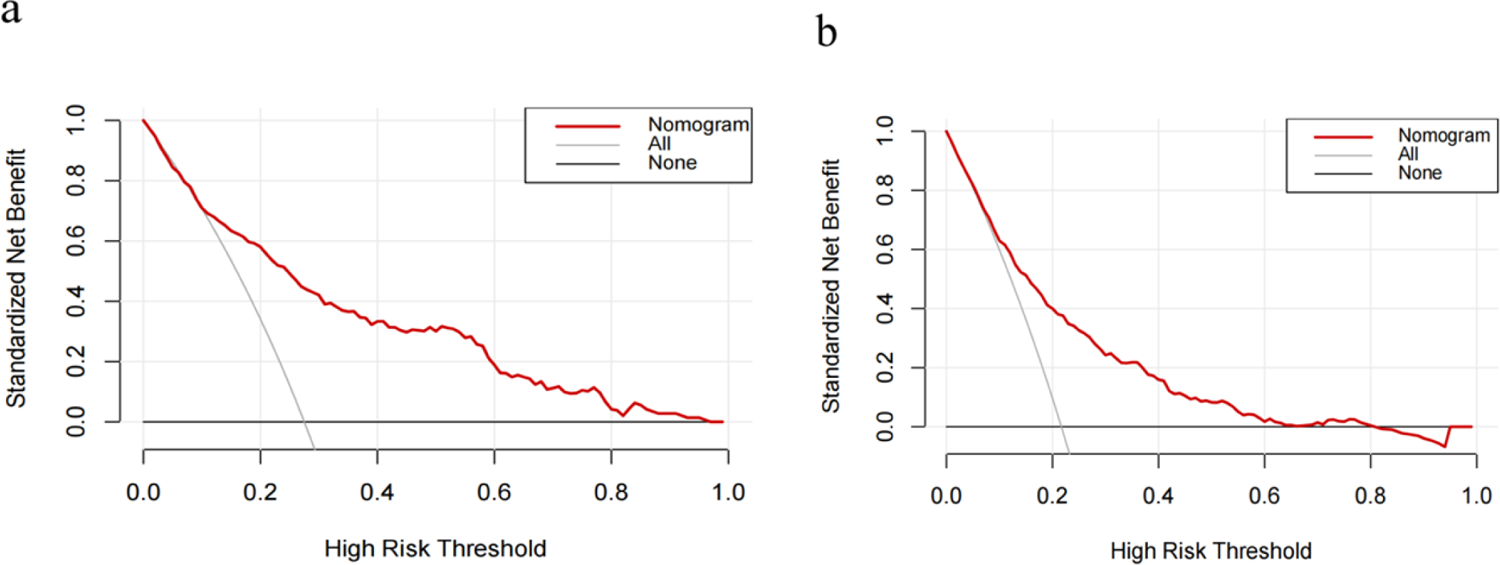
Decision curve analysis (DCA) of the nomogram (a DCA of training set, b DCA of validation set)

**Figure 8.**
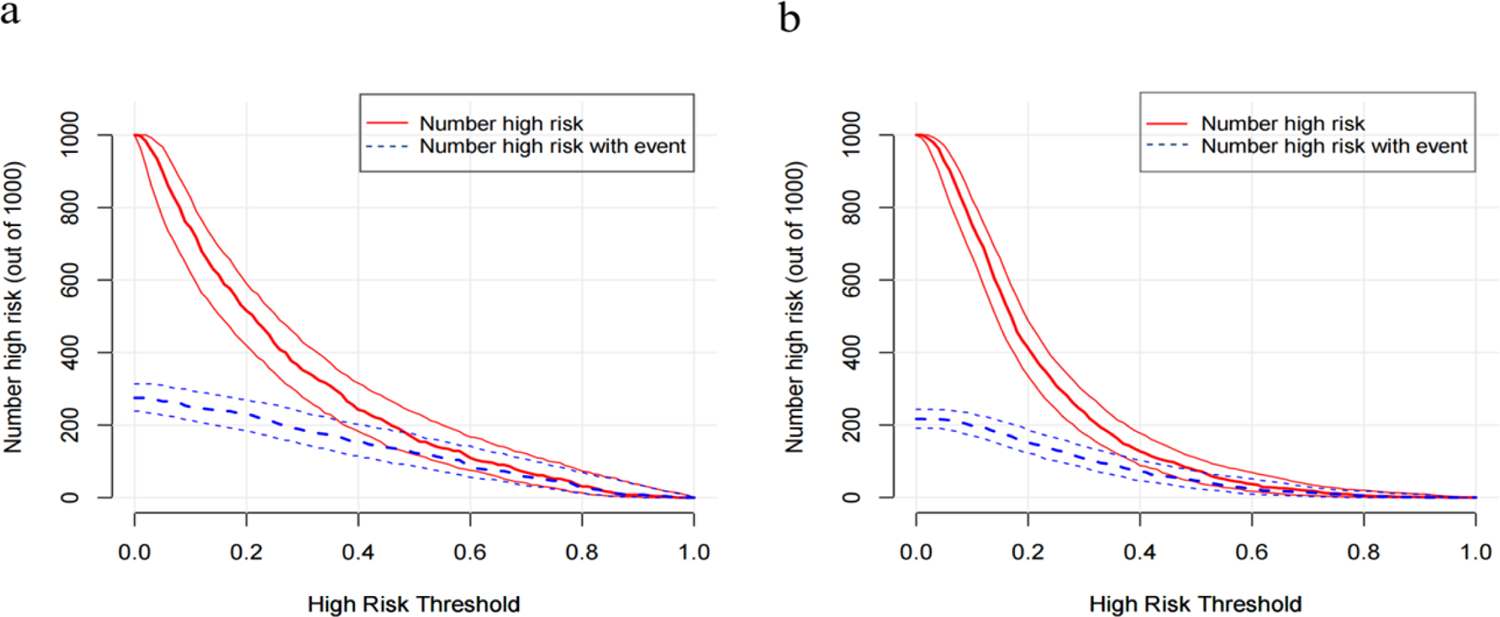
Clinical Impact Curve (CIC) of the nomogram (a CIC of training set, b CIC of validation set)

## Discussion

Based on the increased CHD incidence, its health and economic burdens have increased considerably[20]. Although the prognosis of CHD patients has improved due to expanded PCI use, estimating the long-term risk of MACEs among ACS patients post-PCI is frequently still required due to residual cardiovascular risk [21, 22].

Atherosclerosis is a chronic disease process that dynamically develops in the cardiovascular setting[23]. Several factors related to inflammation, the immune system, and metabolic disorders arising from genetic, environmental, and behavioral drivers accelerate the progression of atherosclerosis, which subsequently contributes to the development of ACS [24–29]. Thus, rather than any single factor, the onset and progression of ACS and its residual risk are influenced by the interaction of multisystem factors.

Several risk and prognosis assessment tools for cardiovascular diseases have been developed to guide clinical practice by identifying individuals at increased risk for MACEs across various populations[30–33]. These tools, along with their corresponding scores, can inform clinical decisions for secondary prevention by identifying high-risk patients who might require ancillary clinical assistance and resources.

However, in addition to some of them requiring complex completion procedures, the clinical validity of these scores is limited. Furthermore, different studies have revealed discrepancies in prognostic judgments for the above scores. For example, GRACE, TIMI, Zwolle, and CADILLAC scores were employed to analyze the 5-year prognosis of STEMI patients post-PCI. Kozieradzka et al. found that for predicting all-cause mortality, the CADILLAC model had the lowest discrimination[14]. After comparing the prognostic accuracy of six scoring models for three-year mortality in STEMI patients, Jarkovsky et al. discovered that longer follow-up periods could best be predicted by GRACE[15]. Scruth et al., on the other hand, concluded that CADILLAC and TIMI scores were better predictors of major cardiac events at one year[16].

Based on the different outcomes and the considerable decision risk, hospitals, particularly county-level hospitals, must urgently establish a clinically simple and effective risk score for evaluating the prognosis of ACS patients post-PCI. Consequently, the new scores will mitigate the risk of adverse cardiovascular events by promoting early and effective measures in high-risk patients post-PCI. Therefore, using routine clinical data from a multicenter, this study constructed a nomogram for long-term risk prediction in ACS patients who underwent PCI. Our findings demonstrated the capacity to build a straightforward model for prognostic evaluation using nine demographic and clinical parameters from existing models created from different databases.

Previous research has linked the clinical profile of ACS patients at admission to prognosis, with age, cardiac insufficiency, and blood pressure clinically recognized as independent markers of poor prognosis following PCI in ACS patients[34–53].

In addition to being the most commonly used measure of left ventricular systolic function, Left Ventricular Ejection Fraction (LVEF), as measured by transthoracic echocardiography, is one of the strongest predictors of MACEs occurrence post-PCI in patients with coronary artery disease. At admission, the LVEF determines the extent of the decline in the left ventricle’s systolic function. Individuals are more likely to experience severe cardiovascular endpoint events due to the long-term decline in cardiac output caused by myocardial infarction.

According to the American Society of Echocardiography and the European Society of Cardiovascular Imaging, specific thresholds of 52% and 54% for men and women, respectively, define an increased risk of left ventricular dysfunction and early death[54]. Tajstra et al. demonstrated that in ischemic heart failure (LVEF ≤35%), patients with chronic, completely occlusive lesions had a worse long-term prognosis[55]. An observational study of 230,464 cases from the British Cardiovascular Intervention Society angioplasty database revealed that compared to patients with preserved LVEF (50%), patients with moderately impaired LV ejection fraction (30%-49%) had a threefold increase in 30-day post-PCI mortality[56]. As revealed by a study summarizing five randomized clinical trials, patients with reduced LVEF <40% or median LVEF 40%-49% had an increased risk of all-cause mortality, cardiac death, and a composite risk of cardiac death in the context of coronary artery disease treated with clinically indicated PCI[57]. In contrast, recent cohort studies have shown inconsistent findings, with LVEF below 60% or over 65% being associated with an increased mortality risk. These findings contradict the previous consensus that associated an increased mortality risk only with severely low levels of LVEF [58]. Furthermore, an Australian study discovered that at higher LVEF levels, women had a greater risk of death[59]. Since these studies were conducted on the general population undergoing echocardiography, the association between LVEF and adverse outcomes in coronary artery disease patients following PCI remains unclear.

Herein, we attempted to examine the connection between LVEF and the risk of MACEs in ACS patients undergoing PCI in a large observational cohort of county-level multicenter hospitals. Using restricted cubic splines, we evaluated a non-linear relationship between LVEF and adverse cardiovascular events. We discovered that compared with patients with intact ejection (>50%), patients with a lower ejection fraction (50%) had a higher incidence of post-PCI MACEs. This result is consistent with the findings of a prior study, which indicated that when LVEF was evaluated using cardiac magnetic resonance, the TIMI risk score had an improved capacity to predict all-cause mortality, reinfarction, and new-onset congestive heart failure within a year following infarction[60].

Additionally, the multivessel disease was found to be an independent predictor of MACEs in patients post-PCI[60, 61]. An increased number of diseased vessels indicates more extensive and complex coronary lesions, necessitating more stents, balloons, and other interventional devices used during PCI, which substantially increase the risk of damage to coronary vessels and cardiomyocytes, and in turn leading to thrombosis and microcirculatory disorders, consequently increasing the risk of post-PCI adverse events[42, 53, 62, 63].

The other independent factor in predicting adverse events in ACS patients was serum homocysteine (Hcy) levels. Elevated serum homocysteine levels can lead to endothelial dysfunction in ACS patients, exacerbating coronary atherosclerosis and producing ischemic cardiovascular events[64, 65].

Following the adjustment for other clinical traits in the study’s participants, we also observed that LDL-C levels remained associated with the risk of adverse events in PCI recipients. This finding stresses the importance of appropriate risk reduction methods as it underlines the large residual risk in these patients despite successful revascularization. Furthermore, a multicenter study investigating the association between LDL-C and long-term cardiovascular events post-PCI linked higher LDL-C levels with an increased risk of late cardiovascular events [66]. Therefore, prompt initiation of intensive statin therapy may provide early clinical benefit after ACS, and long-term adherence to optimal lipid-lowering therapy may effectively reduce long-term cardiovascular events post-PCI [64].

Notably, due to urbanization, significant geographical disparities were observed in the course and prognosis of ACS patients in developing nations, with provincial hospital patients being younger and having lower fatality rates than those treated in district hospitals[67, 68]. Furthermore, the Prospective Urban Rural Epidemiologic (PURE) study revealed that underprivileged populations in low-income countries faced challenges regarding access to primary and secondary prevention[5].

As a quantitative tool for evaluating clinical risk and benefit, our nomogram can facilitate early identification of high-risk individuals by offering primary care practitioners and patients more unbiased and accurate information. Furthermore, it will support clinicians with decision-making on secondary prevention and management strategies, while reducing the unnecessary financial burden of healthcare on patients.

In summary, our nomogram aims to provide novel perspectives for county-level post-PCI rehabilitation programs in LMIC, and consequently lower the incidence of severe adverse cardiovascular events in post-PCI patients.

## Limitations

First, this study created a clinical prediction model by examining independent risk factors for long-term MACEs in PCI patients. Thus, the chosen indicators primarily comprise those commonly employed in clinical contexts.

Second, the study exclusively assessed the prognostic accuracy of the predictive nomogram in ACS patients undergoing PCI. More research is needed to determine whether the predictive nomogram exerts a similar clinical effect on ACS patients undergoing alternative therapies, such as coronary artery bypass graft.

Third, due to the low reliability of telephone follow-up, the study has not distinguished between cardiovascular and non-cardiovascular as well as cancer-specific causes of mortality.

Fourth, since the study was conducted across multiple centers, there were variations in surgical equipment and physician experience. As a result, the PCI outcomes were inevitably impacted, potentially affecting the prognosis of patients. Therefore, the findings of this investigation could be compromised.

## Author Contributions

Conceptualization, Bo Zhou and Yue Lu; Investigation, Yaqian Wang; Methodology, Yaqian Wang and Yue Lu; Formal analysis and Writing-original draft, Yue Lu; All authors approved the final version of the manuscript. All authors have read and agreed to the published version of the manuscript.

## Data Availability

Data supporting results are saved at the server of the First Hospital of China Medical University. Due to patient confidentiality, raw data are not made publicly

## Acknowledgments

We thank all the participants and investigators involved in this study.

## Reference

[1] Global burden of 369 diseases and injuries in 204 countries and territories, 1990-2019: a systematic analysis for the Global Burden of Disease Study 2019. Lancet. 2020;396:1204–22.https://doi.org/10.1016/s0140-6736(20)30925-9.

[2] O’Flaherty M, Allender S, Taylor R, Stevenson C, Peeters A, Capewell S. The decline in coronary heart disease mortality is slowing in young adults (Australia 1976-2006): a time trend analysis. Int J Cardiol. 2012;158:193–8.https://doi.org/10.1016/j.ijcard.2011.01.016.

[3] Ananth CV, Rutherford C, Rosenfeld EB, Brandt JS, Graham H, Kostis WJ, et al. Epidemiologic Trends and Risk Factors Associated with the Decline in Mortality from Coronary Heart Disease in the United States, 1990-2019. Am Heart J. 2023.https://doi.org/10.1016/j.ahj.2023.05.006.

[4] Report on Cardiovascular Health and Diseases in China 2021: An Updated Summary. Biomed Environ Sci. 2022;35:573–603.https://doi.org/10.3967/bes2022.079.

[5] Rosengren A, Smyth A, Rangarajan S, Ramasundarahettige C, Bangdiwala SI, AlHabib KF, et al. Socioeconomic status and risk of cardiovascular disease in 20 low-income, middle-income, and high-income countries: the Prospective Urban Rural Epidemiologic (PURE) study. Lancet Glob Health. 2019;7:e748–e60.https://doi.org/10.1016/s2214-109x(19)30045-2.

[6] Xavier D, Pais P, Devereaux PJ, Xie C, Prabhakaran D, Reddy KS, et al. Treatment and outcomes of acute coronary syndromes in India (CREATE): a prospective analysis of registry data. Lancet. 2008;371:1435–42.https://doi.org/10.1016/s0140-6736(08)60623-6.

[7] Fuster V, Kovacic JC. Acute coronary syndromes: pathology, diagnosis, genetics, prevention, and treatment. Circ Res. 2014;114:1847–51.https://doi.org/10.1161/circresaha.114.302806.

[8] Kumar A, Cannon CP. Acute coronary syndromes: diagnosis and management, part I. Mayo Clin Proc. 2009;84:917–38.https://doi.org/10.1016/s0025-6196(11)60509-0.

[9] Bhatt DL, Lopes RD, Harrington RA. Diagnosis and Treatment of Acute Coronary Syndromes: A Review. Jama. 2022;327:662–75.https://doi.org/10.1001/jama.2022.0358.

[10] Kamran H, Jneid H, Kayani WT, Virani SS, Levine GN, Nambi V, et al. Oral Antiplatelet Therapy After Acute Coronary Syndrome: A Review. Jama. 2021;325:1545–55.https://doi.org/10.1001/jama.2021.0716.

[11] Hoole SP, Bambrough P. Recent advances in percutaneous coronary intervention. Heart. 2020;106:1380–6.https://doi.org/10.1136/heartjnl-2019-315707.

[12] Chacko L, J PH, Rajkumar C, Nowbar AN, Kane C, Mahdi D, et al. Effects of Percutaneous Coronary Intervention on Death and Myocardial Infarction Stratified by Stable and Unstable Coronary Artery Disease: A Meta-Analysis of Randomized Controlled Trials. Circ Cardiovasc Qual Outcomes. 2020;13:e006363.https://doi.org/10.1161/circoutcomes.119.006363.

[13] Zhang S, Wang W, Sawhney JPS, Krittayaphong R, Kim HS, Nhan VT, et al. Antithrombotic management and long-term outcomes following percutaneous coronary intervention for acute coronary syndrome in Asia. Int J Cardiol. 2020;310:16–22.https://doi.org/10.1016/j.ijcard.2020.01.008.

[14] Kozieradzka A, Kamiński KA, Maciorkowska D, Olszewska M, Dobrzycki S, Nowak K, et al. GRACE, TIMI, Zwolle and CADILLAC risk scores--do they predict 5-year outcomes after ST-elevation myocardial infarction treated invasively? Int J Cardiol. 2011;148:70–5.https://doi.org/10.1016/j.ijcard.2009.10.026.

[15] Littnerova S, Kala P, Jarkovsky J, Kubkova L, Prymusova K, Kubena P, et al. GRACE Score among Six Risk Scoring Systems (CADILLAC, PAMI, TIMI, Dynamic TIMI, Zwolle) Demonstrated the Best Predictive Value for Prediction of Long-Term Mortality in Patients with ST-Elevation Myocardial Infarction. PLoS One. 2015;10:e0123215.https://doi.org/10.1371/journal.pone.0123215.

[16] Scruth EA, Cheng E, Worrall-Carter L. Risk score comparison of outcomes in patients presenting with ST-elevation myocardial infarction treated with percutaneous coronary intervention. Eur J Cardiovasc Nurs. 2013;12:330–6.https://doi.org/10.1177/1474515112449412.

[17] Zhu KF, Wang YM, Zhu JZ, Zhou QY, Wang NF. National prevalence of coronary heart disease and its relationship with human development index: A systematic review. Eur J Prev Cardiol. 2016;23:530–43.https://doi.org/10.1177/2047487315587402.

[18] Zhong Q, Gao Y, Zheng X, Chen J, Masoudi FA, Lu Y, et al. Geographic Variation in Process and Outcomes of Care for Patients With Acute Myocardial Infarction in China From 2001 to 2015. JAMA Netw Open. 2020;3:e2021182.https://doi.org/10.1001/jamanetworkopen.2020.21182.

[19] Collins GS, Reitsma JB, Altman DG, Moons KG. Transparent Reporting of a multivariable prediction model for Individual Prognosis or Diagnosis (TRIPOD): the TRIPOD statement. Ann Intern Med. 2015;162:55–63.https://doi.org/10.7326/m14-0697.

[20] Zheng X, Curtis JP, Hu S, Wang Y, Yang Y, Masoudi FA, et al. Coronary Catheterization and Percutaneous Coronary Intervention in China: 10-Year Results From the China PEACE-Retrospective CathPCI Study. JAMA Intern Med. 2016;176:512–21.https://doi.org/10.1001/jamainternmed.2016.0166.

[21] Ajala ON, Everett BM. Targeting Inflammation to Reduce Residual Cardiovascular Risk. Curr Atheroscler Rep. 2020;22:66.https://doi.org/10.1007/s11883-020-00883-3.

[22] Hoogeveen RC, Ballantyne CM. Residual Cardiovascular Risk at Low LDL: Remnants, Lipoprotein(a), and Inflammation. Clin Chem. 2021;67:143–53.https://doi.org/10.1093/clinchem/hvaa252.

[23] Libby P, Buring JE, Badimon L, Hansson GK, Deanfield J, Bittencourt MS, et al. Atherosclerosis. Nat Rev Dis Primers. 2019;5:56.https://doi.org/10.1038/s41572-019-0106-z.

[24] Wolf D, Ley K. Immunity and Inflammation in Atherosclerosis. Circ Res. 2019;124:315–27.https://doi.org/10.1161/circresaha.118.313591.

[25] Poznyak A, Grechko AV, Poggio P, Myasoedova VA, Alfieri V, Orekhov AN. The Diabetes Mellitus-Atherosclerosis Connection: The Role of Lipid and Glucose Metabolism and Chronic Inflammation. Int J Mol Sci. 2020;21.https://doi.org/10.3390/ijms21051835.

[26] Ridker PM, Everett BM, Thuren T, MacFadyen JG, Chang WH, Ballantyne C, et al. Antiinflammatory Therapy with Canakinumab for Atherosclerotic Disease. N Engl J Med. 2017;377:1119–31.https://doi.org/10.1056/NEJMoa1707914.

[27] Gallone G, Baldetti L, Pagnesi M, Latib A, Colombo A, Libby P, et al. Medical Therapy for Long-Term Prevention of Atherothrombosis Following an Acute Coronary Syndrome: JACC State-of-the-Art Review. J Am Coll Cardiol. 2018;72:2886–903.https://doi.org/10.1016/j.jacc.2018.09.052.

[28] Eikelboom JW, Connolly SJ, Bosch J, Dagenais GR, Hart RG, Shestakovska O, et al. Rivaroxaban with or without Aspirin in Stable Cardiovascular Disease. N Engl J Med. 2017;377:1319–30.https://doi.org/10.1056/NEJMoa1709118.

[29] Mauri L, Kereiakes DJ, Yeh RW, Driscoll-Shempp P, Cutlip DE, Steg PG, et al. Twelve or 30 months of dual antiplatelet therapy after drug-eluting stents. N Engl J Med. 2014;371:2155–66.https://doi.org/10.1056/NEJMoa1409312.

[30] Fox KA, Dabbous OH, Goldberg RJ, Pieper KS, Eagle KA, Van de Werf F, et al. Prediction of risk of death and myocardial infarction in the six months after presentation with acute coronary syndrome: prospective multinational observational study (GRACE). Bmj. 2006;333:1091.https://doi.org/10.1136/bmj.38985.646481.55.

[31] Halkin A, Singh M, Nikolsky E, Grines CL, Tcheng JE, Garcia E, et al. Prediction of mortality after primary percutaneous coronary intervention for acute myocardial infarction: the CADILLAC risk score. J Am Coll Cardiol. 2005;45:1397–405.https://doi.org/10.1016/j.jacc.2005.01.041.

[32] Antman EM, Cohen M, Bernink PJ, McCabe CH, Horacek T, Papuchis G, et al. The TIMI risk score for unstable angina/non-ST elevation MI: A method for prognostication and therapeutic decision making. Jama. 2000;284:835–42.https://doi.org/10.1001/jama.284.7.835.

[33] De Luca G, Suryapranata H, van’t Hof AW, de Boer MJ, Hoorntje JC, Dambrink JH, et al. Prognostic assessment of patients with acute myocardial infarction treated with primary angioplasty: implications for early discharge. Circulation. 2004;109:2737–43.https://doi.org/10.1161/01.Cir.0000131765.73959.87.

[34] Kong S, Chen C, Zheng G, Yao H, Li J, Ye H, et al. A prognostic nomogram for long-term major adverse cardiovascular events in patients with acute coronary syndrome after percutaneous coronary intervention. BMC Cardiovasc Disord. 2021;21:253.https://doi.org/10.1186/s12872-021-02051-0.

[35] Pedersen F, Butrymovich V, Kelbæk H, Wachtell K, Helqvist S, Kastrup J, et al. Short- and long-term cause of death in patients treated with primary PCI for STEMI. J Am Coll Cardiol. 2014;64:2101–8.https://doi.org/10.1016/j.jacc.2014.08.037.

[36] Yamaji K, Shiomi H, Morimoto T, Nakatsuma K, Toyota T, Ono K, et al. Effects of Age and Sex on Clinical Outcomes After Percutaneous Coronary Intervention Relative to Coronary Artery Bypass Grafting in Patients With Triple-Vessel Coronary Artery Disease. Circulation. 2016;133:1878–91.https://doi.org/10.1161/circulationaha.115.020955.

[37] Giustino G, Redfors B, Brener SJ, Kirtane AJ, Généreux P, Maehara A, et al. Correlates and prognostic impact of new-onset heart failure after ST-segment elevation myocardial infarction treated with primary percutaneous coronary intervention: insights from the INFUSE-AMI trial. Eur Heart J Acute Cardiovasc Care. 2018;7:339–47.https://doi.org/10.1177/2048872617719649.

[38] Hwang D, Lee JM, Yang S, Chang M, Zhang J, Choi KH, et al. Role of Post-Stent Physiological Assessment in a Risk Prediction Model After Coronary Stent Implantation. JACC Cardiovasc Interv. 2020;13:1639–50.https://doi.org/10.1016/j.jcin.2020.04.041.

[39] Qin Y, Wei X, Han H, Wen Y, Gu K, Ruan Y, et al. Association between age and readmission after percutaneous coronary intervention for acute myocardial infarction. Heart. 2020;106:1595–603.https://doi.org/10.1136/heartjnl-2019-316103.

[40] Ye Z, Xu Y, Tang L, Wu M, Wu B, Zhu T, et al. Predicting long-term prognosis after percutaneous coronary intervention in patients with new onset ST-elevation myocardial infarction: development and external validation of a nomogram model. Cardiovasc Diabetol. 2023;22:87.https://doi.org/10.1186/s12933-023-01820-9.

[41] Fang C, Chen Z, Zhang J, Jin X, Yang M. Construction and evaluation of nomogram model for individualized prediction of risk of major adverse cardiovascular events during hospitalization after percutaneous coronary intervention in patients with acute ST-segment elevation myocardial infarction. Front Cardiovasc Med. 2022;9:1050785.https://doi.org/10.3389/fcvm.2022.1050785.

[42] Liu HW, Han YL, Jin QM, Wang XZ, Ma YY, Wang G, et al. One-year Outcomes in Patients with ST-segment Elevation Myocardial Infarction Caused by Unprotected Left Main Coronary Artery Occlusion Treated by Primary Percutaneous Coronary Intervention. Chin Med J (Engl). 2018;131:1412–9.https://doi.org/10.4103/0366-6999.233948.

[43] Qin Z, Du Y, Zhou Q, Lu X, Luo L, Zhang Z, et al. NT-proBNP and Major Adverse Cardiovascular Events in Patients with ST-Segment Elevation Myocardial Infarction Who Received Primary Percutaneous Coronary Intervention: A Prospective Cohort Study. Cardiol Res Pract. 2021;2021:9943668.https://doi.org/10.1155/2021/9943668.

[44] Song JJ, Liu YP, Wang WY, Yang J, Wen J, Chen J, et al. Development and validation of a nomogram predicting one-year mortality in patients undergoing percutaneous coronary intervention. J Geriatr Cardiol. 2022;19:960–9.https://doi.org/10.11909/j.issn.1671-5411.2022.12.003.

[45] Song J, Liu Y, Wang W, Chen J, Yang J, Wen J, et al. A nomogram predicting 30-day mortality in patients undergoing percutaneous coronary intervention. Front Cardiovasc Med. 2022;9:897020.https://doi.org/10.3389/fcvm.2022.897020.

[46] Hua C, Tian H, Wang Y, Zheng J, Liu P, Zhang B, et al. Development and Validation of a Nomogram for Predicting Mortality in Patients with Atrial Fibrillation and Acute Coronary Syndrome Who Underwent Percutaneous Coronary Intervention in a Chinese Multicenter Cohort. Appl Bionics Biomech. 2022;2022:2586400.https://doi.org/10.1155/2022/2586400.

[47] Bo X, Liu Y, Yang M, Lu Z, Zhao Y, Chen L. Development and Validation of a Nomogram of In-hospital Major Adverse Cardiovascular and Cerebrovascular Events in Patients With Acute Coronary Syndrome. Front Cardiovasc Med. 2021;8:699023.https://doi.org/10.3389/fcvm.2021.699023.

[48] Zhao E, Xie H, Zhang Y. A Nomogram Based on Apelin-12 for the Prediction of Major Adverse Cardiovascular Events after Percutaneous Coronary Intervention among Patients with ST-Segment Elevation Myocardial Infarction. Cardiovasc Ther. 2020;2020:9416803.https://doi.org/10.1155/2020/9416803.

[49] Wang Y, Wang W, Jia S, Gao M, Zheng S, Wang J, et al. Development of a nomogram for the prediction of in-hospital mortality in patients with acute ST-elevation myocardial infarction after primary percutaneous coronary intervention: a multicentre, retrospective, observational study in Hebei province, China. BMJ Open. 2022;12:e056101.https://doi.org/10.1136/bmjopen-2021-056101.

[50] Pan D, Xiao S, Hu Y, Pan Q, Wu Q, Wang X, et al. Clinical Nomogram to Predict Major Adverse Cardiac Events in Acute Myocardial Infarction Patients within 1 Year of Percutaneous Coronary Intervention. Cardiovasc Ther. 2021;2021:3758320.https://doi.org/10.1155/2021/3758320.

[51] Zheng YY, Wu TT, Gao Y, Guo QQ, Ma YY, Zhang JC, et al. A Novel ABC Score Predicts Mortality in Non-ST-Segment Elevation Acute Coronary Syndrome Patients Who underwent Percutaneous Coronary Intervention. Thromb Haemost. 2021;121:297–308.https://doi.org/10.1055/s-0040-1718411.

[52] Zhao X, Yuan X, Zhong Y, Wang G. Risk Factor Analysis and Construction of a Prognostic Model after Percutaneous Coronary Intervention in Patients with Chronic Total Occlusion. Comput Math Methods Med. 2022;2022:9902380.https://doi.org/10.1155/2022/9902380.

[53] Li M, Hou J, Gu X, Weng R, Zhong Z, Liu S. Incidence and risk factors of in-stent restenosis after percutaneous coronary intervention in patients from southern China. Eur J Med Res. 2022;27:12.https://doi.org/10.1186/s40001-022-00640-z.

[54] Lang RM, Badano LP, Mor-Avi V, Afilalo J, Armstrong A, Ernande L, et al. Recommendations for cardiac chamber quantification by echocardiography in adults: an update from the American Society of Echocardiography and the European Association of Cardiovascular Imaging. Eur Heart J Cardiovasc Imaging. 2015;16:233–70.https://doi.org/10.1093/ehjci/jev014.

[55] Tajstra M, Pyka Ł, Gorol J, Pres D, Gierlotka M, Gadula-Gacek E, et al. Impact of Chronic Total Occlusion of the Coronary Artery on Long-Term Prognosis in Patients With Ischemic Systolic Heart Failure: Insights From the COMMIT-HF Registry. JACC Cardiovasc Interv. 2016;9:1790–7.https://doi.org/10.1016/j.jcin.2016.06.007.

[56] Almudarra SS, Gale CP, Baxter PD, Fleming SJ, Brogan RA, Ludman PF, et al. Comparative outcomes after unprotected left main stem percutaneous coronary intervention: a national linked cohort study of 5,065 acute and elective cases from the BCIS Registry (British Cardiovascular Intervention Society). JACC Cardiovasc Interv. 2014;7:717–30.https://doi.org/10.1016/j.jcin.2014.03.005.

[57] Siontis GC, Branca M, Serruys P, Silber S, Räber L, Pilgrim T, et al. Impact of left ventricular function on clinical outcomes among patients with coronary artery disease. Eur J Prev Cardiol. 2019;26:1273–84.https://doi.org/10.1177/2047487319841939.

[58] Wehner GJ, Jing L, Haggerty CM, Suever JD, Leader JB, Hartzel DN, et al. Routinely reported ejection fraction and mortality in clinical practice: where does the nadir of risk lie? Eur Heart J. 2020;41:1249–57.https://doi.org/10.1093/eurheartj/ehz550.

[59] Stewart S, Playford D, Scalia GM, Currie P, Celermajer DS, Prior D, et al. Ejection fraction and mortality: a nationwide register-based cohort study of 499 153 women and men. Eur J Heart Fail. 2021;23:406–16.https://doi.org/10.1002/ejhf.2047.

[60] Eitel I, de Waha S, Wöhrle J, Fuernau G, Lurz P, Pauschinger M, et al. Comprehensive prognosis assessment by CMR imaging after ST-segment elevation myocardial infarction. J Am Coll Cardiol. 2014;64:1217–26.https://doi.org/10.1016/j.jacc.2014.06.1194.

[61] Liu KL, Lin SM, Chang CH, Chen YC, Chu PH. Plasma angiopoietin-1 level, left ventricular ejection fraction, and multivessel disease predict development of 1-year major adverse cardiovascular events in patients with acute ST elevation myocardial infarction - a pilot study. Int J Cardiol. 2015;182:155–60.https://doi.org/10.1016/j.ijcard.2014.12.172.

[62] Cho YK, Nam CW. Percutaneous coronary intervention in patients with multi-vessel coronary artery disease: a focus on physiology. Korean J Intern Med. 2018;33:851–9.https://doi.org/10.3904/kjim.2018.006.

[63] Denktas AE, Paniagua D, Jneid H. Coronary Physiology Assessment for the Diagnosis and Treatment of Stable Ischemic Heart Disease. Curr Atheroscler Rep. 2016;18:62.https://doi.org/10.1007/s11883-016-0613-2.

[64] Koskinas KC, Mach F, Räber L. Lipid-lowering therapy and percutaneous coronary interventions. EuroIntervention. 2021;16:1389–403.https://doi.org/10.4244/eij-d-20-00999.

[65] Fu Z, Qian G, Xue H, Guo J, Chen L, Yang X, et al. Hyperhomocysteinemia is an independent predictor of long-term clinical outcomes in Chinese octogenarians with acute coronary syndrome. Clin Interv Aging. 2015;10:1467–74.https://doi.org/10.2147/cia.S91652.

[66] Sud M, Han L, Koh M, Abdel-Qadir H, Austin PC, Farkouh ME, et al. Low-Density Lipoprotein Cholesterol and Adverse Cardiovascular Events After Percutaneous Coronary Intervention. J Am Coll Cardiol. 2020;76:1440–50.https://doi.org/10.1016/j.jacc.2020.07.033.

[67] Xu H, Yang Y, Wang C, Yang J, Li W, Zhang X, et al. Association of Hospital-Level Differences in Care With Outcomes Among Patients With Acute ST-Segment Elevation Myocardial Infarction in China. JAMA Netw Open. 2020;3:e2021677.https://doi.org/10.1001/jamanetworkopen.2020.21677.

[68] Henry TD, Jollis JG. Lessons Learned From Acute Myocardial Infarction Care in China. JAMA Netw Open. 2020;3:e2021768.https://doi.org/10.1001/jamanetworkopen.2020.21768.

